# Caregiving in a Tertiary Health Institution in North Central Nigeria: Support Types and Financial Burden

**DOI:** 10.1101/2022.07.04.22276588

**Authors:** Oluwatosin Wuraola Akande, Mojirola Martina Fasiku, Oladimeji Akeem Bolarinwa, Tanimola Makanjuola Akande

**Affiliations:** Department of Epidemiology and Community Health, University of Ilorin Teaching Hospital, Ilorin, Kwara State, Nigeria

**Keywords:** Caregivers, patient admission, Nigeria

## Abstract

**Background:** Caregivers play an important role in informal patient management. Identification of the support types and the challenges faced by caregivers will provide information on strategies to ease this burden. This study aimed to describe the support types and financial burden among caregivers in a tertiary hospital in North Central Nigeria.

**Methods:** This was a cross-sectional study conducted among caregivers of in-patients admitted in a tertiary hospital in North Central Nigeria. Data was collected using a pre-tested interviewer-administered questionnaire. Data was analysed using the Statistical Package for Social Sciences package version 23. Results were reported in frequencies and proportions, and presented in prose, tables and charts.

**Results:** A total of 400 caregivers were recruited. Mean age was 38.32 ± 12.82 years and most (66.0%) were females. Caregivers supported their patients by running errands (96.3%) and 85.3% reported caregiving as stressful. The reported errands were purchase of medications (92.3%), supply of non-medical needs (63.3%), submission of laboratory samples and collection of results (52.3%) and service payment (47.5%). About two thirds (63.2%) reported loss of income while caregiving and about half (50.8%) provided financial support to the patients.

**Conclusion:** This study suggests that majority of caregivers experience significant physical and financial burden while caregiving. This burden can be eased off by the simplification of payment and laboratory processes, and employment of more staff to support patients admitted in the wards. The financial burden experienced by caregivers reinforces the need to encourage more Nigerians to enrol in a health insurance scheme.

## Introduction

A caregiver is an unpaid person usually a family member or a friend who is involved in providing care whether formal or informal, full time or part time, to a patient in their home or a care setting.[1]The need for this social support system in care settings such as the hospital is recognised, where these informal care givers provide a wide array of support to their patients.[1]This support could be physical (assisting in carrying out activities of daily living or running errands), financial, emotional or social.[1,2]

The role caregivers play in patient management has been identified and the burden from this role is recognised.[3,4] Caregivers provide care to their patients in form of assisting with activities of daily such as bathing, feeding, helping with urination and defaecation.[5] They also perform some nursing duties including serving medication and turning patients in bed.[5]Caregiver burden has been linked to a negative impact on their physical, psychological and emotional health.[4]

Caregivers in low resource settings are particularly more burdened in this regard compared to those in developed countries.[5,6]Healthcare workers in these settings are often over-burdened and may shift some of their responsibilities to the informal caregivers.[7,8]In addition, patients and caregivers experience economic burden especially in health systems where out of pocket payment for healthcare is prevalent.[9,10]For example, in Nigeria, the burden faced by caregivers has been associated with the poor economic conditions in the country and poor institutional support especially in public hospitals.[6,7]

An identification of the support types and the challenges faced by caregivers will provide information on strategies to reduce the burden faced by caregivers, especially in low income settings where evidence shows that they are more involved in providing support to their patient while on hospital admission.[5] Extensive electronic literature review has revealed sparse literature regarding the types of support and financial challenges that caregivers experience in Nigeria. This study describes the support types and financial challenges among caregivers in a tertiary hospital in North Central Nigeria. Findings from this study will be useful for the policy makers, hospital administrators and other stakeholders to better manage supportive caregiving services in the hospital.

## Methods

### Study design

This was a hospital-based cross-sectional study conducted to describe the support types and financial challenges experienced by caregivers in-patients in University of Ilorin Teaching Hospital (UITH), Ilorin.

### Study setting

This study was conducted in the University of Ilorin Teaching Hospital (UITH), Ilorin, Kwara State. It is a public owned tertiary hospital located in North Central Nigeria. It is a 650-bedded hospital with an average of about 1,000 in-patient admissions per month. In addition, to rendering its services to patients in Kwara State, patients from surrounding North-central and part of south-western States are also referred to the hospital.

Caregivers play a role in the care of their patients in the hospital, similar to other hospitals in Nigeria.[11]They are primarily responsible for running some errands including payment of bills, and purchase of medications and consumables for their patients. On observation, the hospital provides roofed open spaces around the hospital wards. In addition, a private guest house with 10 rooms is available for caregivers to lodge within the hospital premises. Many caregivers are seen to hang around the corridors close to their patient’s ward during the day and also at night. There is an open local market in the hospital where food and household items can be purchased.

### Study population

Adult caregivers of in-patients in UITH, Ilorin

### Inclusion criteria

Adult caregivers of in-patients who had spent at least one night in UITH, Ilorin.

### Exclusion criteria

Caregivers less than 18 years of age and family/friends on visitation to the hospital.

### Variables

Variables included sociodemographic characteristics (age, gender, religion, ethnicity, marital status, type of family, level of education, employment status and place of residence), relationship with patient, timeline of hospital stay, and perception of care received. Data was also collected on the estimated amount caregivers spend on their patients and their sources of support, and the type of caregiving support they provide.

## Data resource and measurement

### Data collection tool

Data was collected using an interviewer administered questionnaire with close-ended items. The questionnaire was developed from review of relevant literature by the researchers. It was further validated by pretesting for the content validation among 40 respondents in a General Hospital Ilorin (a state government owned tertiary health facility). It was also reviewed by experts for face validity.

### Data collection

Data was collected from participants between 26^th^ September 2019 and 25^th^ October 2019. Demographic variables such as age, gender, religion, level of education, and employment status were collected. The data collection also collected data on variables related to hospital stay of caregivers and perception of caregiving, type of support provided, caregivers’ finances and sources of support. These variables were based on self-reports, which may cause recall bias. However, we tried to reduce this bias to limiting the participants to those who had patients in the ward as at the time of data collection and in the case where a patient had two or more caregivers, the caregiver who had spent more time with the patient was selected.

### Sample Size Determination

Using Leslie Fischer’s Formula for the calculation of sample size for a population less than 10,000,[12] after adjusting for non-response, a sample size of 400 caregivers the calculated minimum sample size was 270. However, a total of 400 caregivers were recruited into the study over a period of one month.

### Sampling method

Caregivers were identified through their patients on admission. They were recruited from all the wards in the hospital. Using the in-patients register in each ward as a sample frame, the patients’ caregivers were proportionally recruited. In each ward, patients randomly selected from the in-patient register based on the allocated number per ward. Caregivers of the selected patients were traced and recruited into the study. In cases where the caregiver did not meet the eligibility criteria, the next patient’s caregiver on the register was recruited. Where the patient had more than one caregiver at the time of data collection, the caregiver who had spent more time with the patient was recruited.

### Data analysis

Data was analysed using the Statistical Package for Social Sciences package version 23. Descriptive statistics (means, standard deviation and percentages) were used to analyze the data. Data was presented in tables (in frequency and proportion) and charts (bar).

### Ethical consideration

Ethical approval was sought from the Ethical Review Committee of UITH, Ilorin. The purpose and benefits of the study was explained to all the study participants after which a written informed consent which was signed or thumb printed was obtained from all. They were assured of the confidentiality and privacy of their information by avoiding the use of identifiers and ensuring data security. All the principles of ethics were adhered to all through the study.

## Results

The majority (35.0%) of the respondents were between 30 and 39 years with mean age of 38.32 ± 12.82 years (Table 1). About two thirds (66%) of the caregivers were females and majority (69.2%) were Muslims. Almost all (89.8%) of the respondents were of Yoruba ethnic group and most of them (81.7%) were married. More than one third (35.4%) attained tertiary education level, while only a minority (16.0%) had no formal education. More than two-third of the respondents (69.2%) were employed, out of which almost two thirds (63.2%) were self-employed. Trading was the most common occupation, accounting for 44.2% of all occupations. Almost three quarters (73.0%) of the respondents resided in the study area. Majority of the caregivers were immediate family members of the patients, with parents, children, siblings and spouses accounting for 35.5%, 18.8%, 13.8%, 11.0% and 15.8% respectively. Less than one fifth (15.8%) were other relatives and less than one tenth (5.1%) were non-relatives.

**Table 1:** Socio-demographic Variables of Caregivers

| **Variables** | **Frequency (%)** |
| --- | --- |
| **Age groups** |  |
| < 20 | 9 (2.3) |
| 20 – 29 | 91 (22.7) |
| 30 – 39 | 140 (35.0) |
| 40 – 49 | 82 (20.5) |
| 50 – 59 | 44 (11.0) |
| ≥ 60 – 69 | 34 (8.5) |
| **Mean ± SD** | 38.32 ± 12.82 |
| **Gender** |  |
| Male | 136 (34.0) |
| Female | 264 (66.0) |
| **Religion** |  |
| Christianity | 123 (30.8) |
| Islam | 277 (69.2) |
| **Ethnicity** |  |
| Yoruba | 359 (89.8) |
| Igbo | 17 (4.3) |
| Hausa | 6 (1.5) |
| Others | 18 (4.4) |
| **Marital Status** |  |
| Married | 327 (81.7) |
| Single | 65 (16.3) |
| Widowed | 8 (2.0) |
| **Type of family** |  |
| Monogamy | 303 (75.7) |
| Polygamy | 97 (24.3) |
| **Level of education** |  |
| No formal education | 64 (16.0) |
| Primary education | 85 (21.3) |
| Secondary education | 105 (26.3) |
| Tertiary education | 142 (35.4) |
| Quranic education | 4 (1.0) |
| **Employment status** |  |
| Employed | 277 (69.2) |
| Unemployed | 108 (27.0) |
| Retiree | 15 (3.8) |
| **Nature of employment** | n=277 |
| Employee | 102 (36.8) |
| Self employed | 175 (63.2) |
| **Main occupation** |  |
| Farming | 24 (8.6) |
| Trading | 123 (44.2) |
| Civil servant | 54 (13.5) |
| Private establishment | 19 (6.8) |
| Artisan | 33 (11.9) |
| Others | 24 (9.0) |
| **Residence** |  |
| Within Ilorin | 292 (73.0) |
| Outside Ilorin | 108 (27.0) |
| **Relationship with patient** |  |
| Parent | 142 (35.5) |
| Sibling | 55 (13.8) |
| Spouse | 44 (11.0) |
| Relative | 63 (15.8) |
| Child | 75 (18.8) |
| Others (friends, domestic aide, etc) | 21 (5.1) |

Running of errands (96.3%) is the commonest support type of caregiving provided to patients on admission in the hospital (Figure 1). Keeping patients’ company (50.5%), feeding patients (37%) and serving medication (28.8%) were also reported as common caregiving roles. Some of the other identified support roles were helping with defecation (25.8%), lifting patients (21%), bathing patients (20.3%) and walking patients (7.8%).

**Figure 1:**
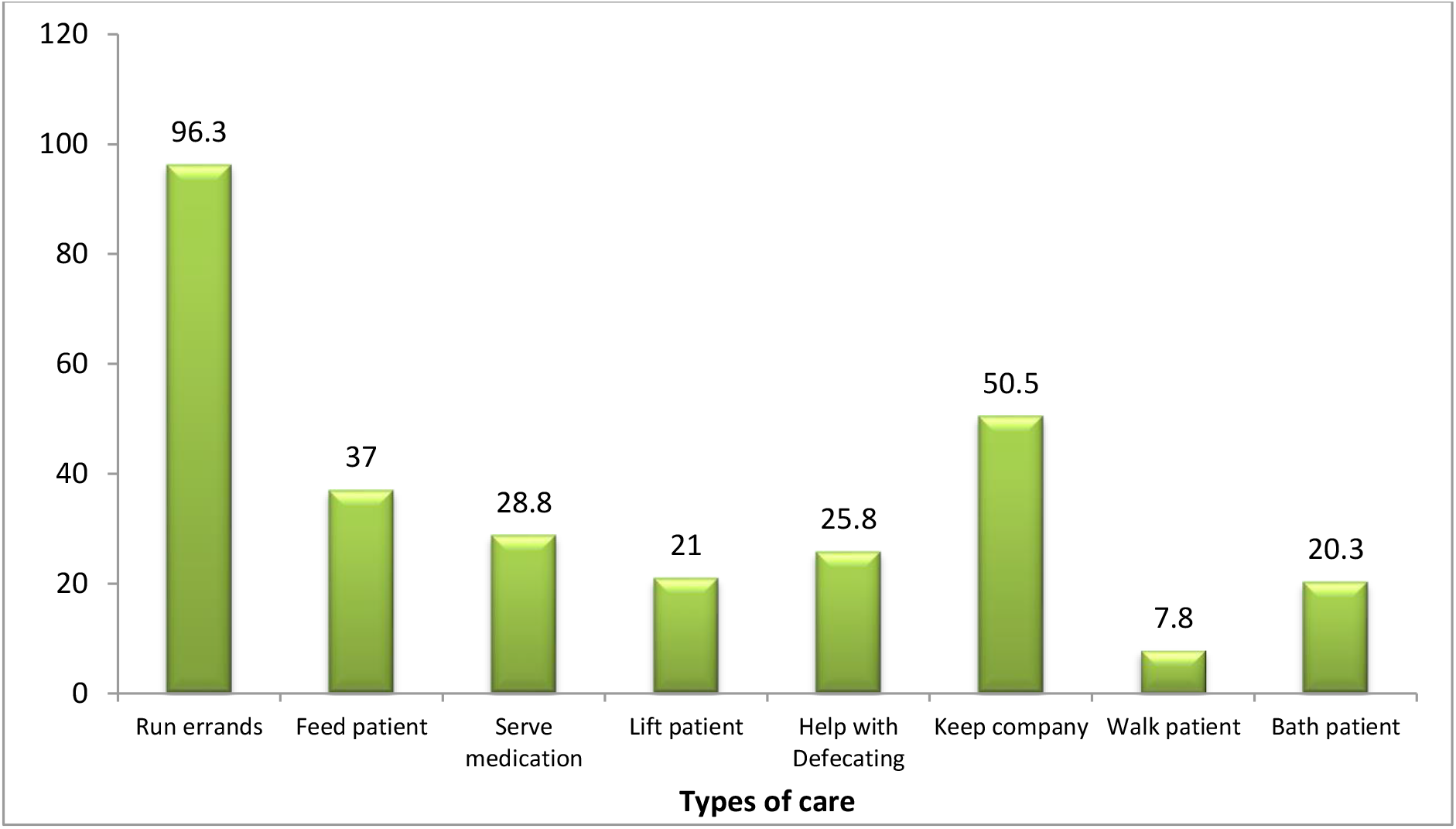
Types of Caregiving Support Provided to Patients *Multiple responses

**Figure 2:**
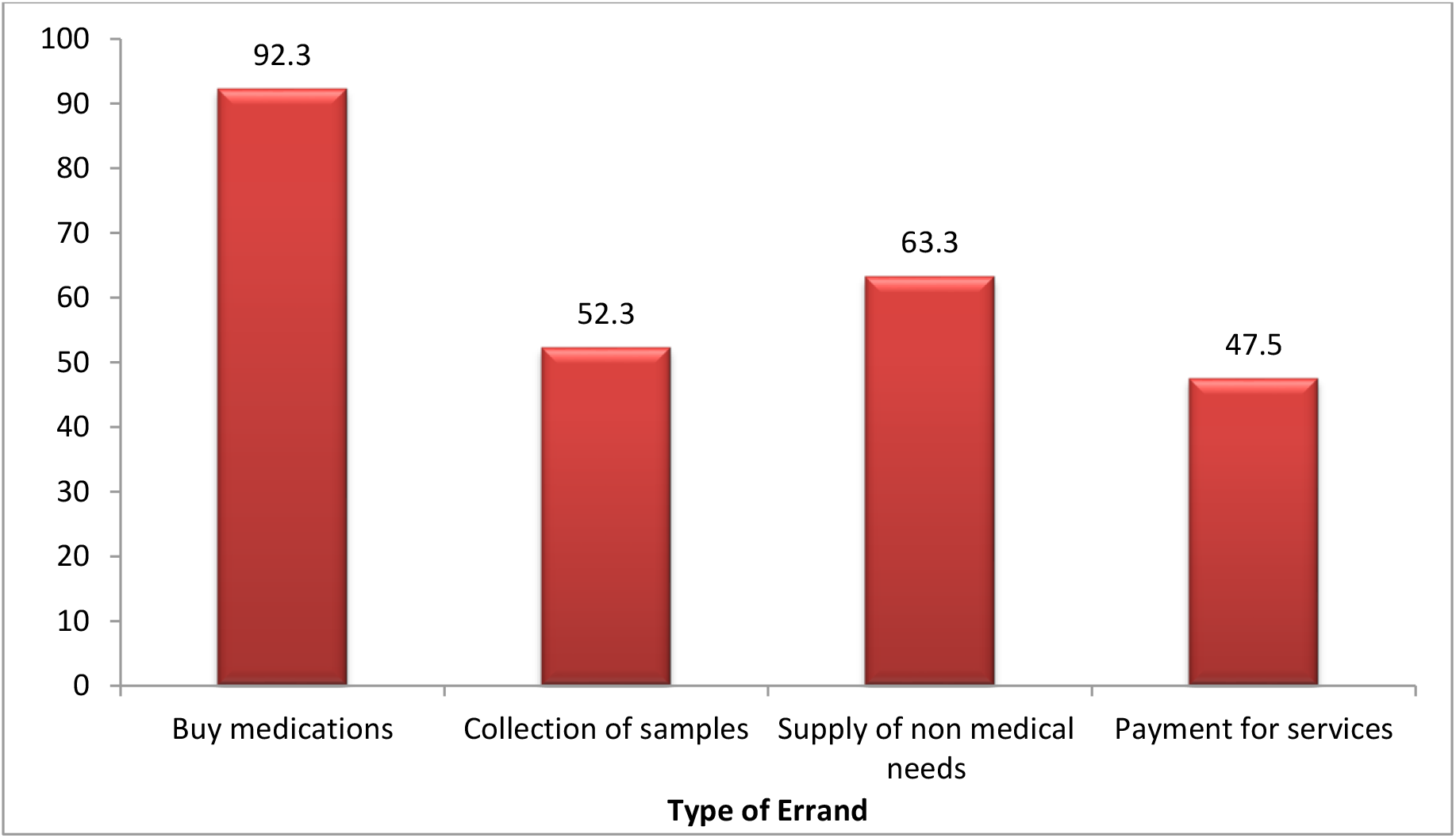
Common Errand Types Provided by the Caregivers *Multiple responses

Almost all (92.3%) of the caregivers run errands to buy medications for the patients, about two thirds (63.3%) supplied non-medical needs, more than half (52.3%) collected samples and almost half (47.5%) of them reportedly made payment for services.

Close to half (47%) of the respondents spent less than one week (between 1-6 days) in the hospital (Table 2). Those who spent more than two weeks and those who spent between one and weeks accounted for 28.5% and 24.5% of respondents respectively. The mean and median length of stay were 1.8 and 2 weeks correspondingly. More than three quarters (79.0%) stayed with their patients both night and day. Less than one fifth (17.7%) stayed only during the day and only a minority (3.3%) stayed only at night.

**Table 2:**
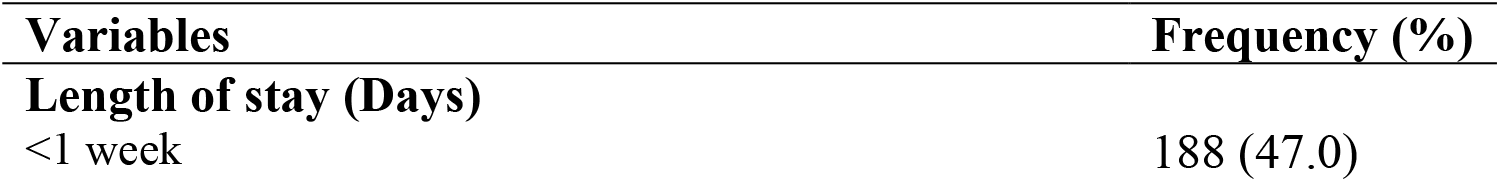
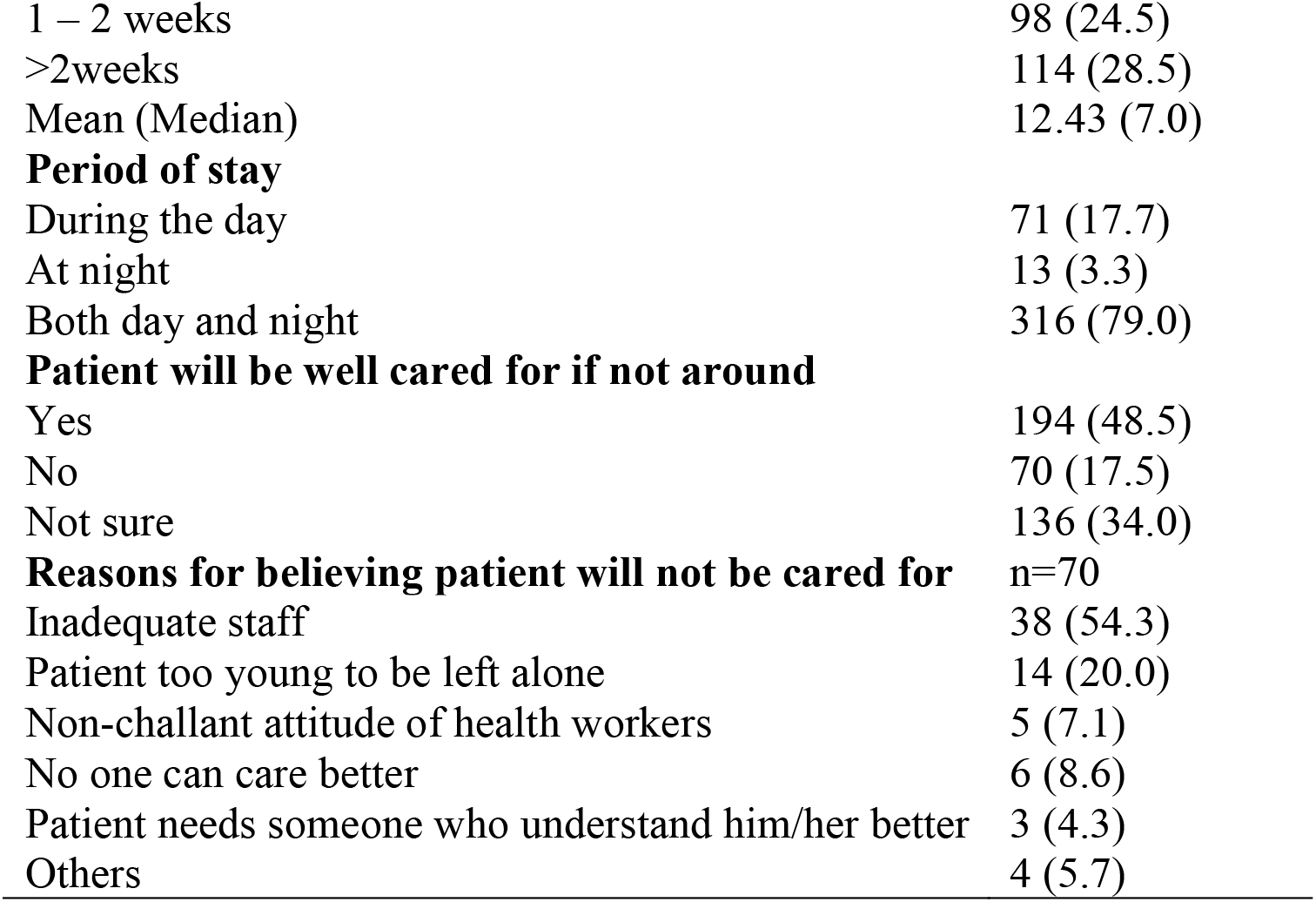
Hospital Stay of Caregivers and Perception of Caregivers towards Caregiving

Majority of the caregivers (88.3%) were aware of the ailment that their patients were being managed for. More than four fifths (85.3%) felt that staying in the hospital had created additional stress for them. Almost half (48.5%) believed their patient will not be well cared for in their absence, another 34.0% were not sure if their patients will be well cared for in their absence and only 17.5%believed their patients will be well cared for in their absence. The most common reasons why respondents believed their patients will not be well cared were inadequate staffing (54.3%), patient being too young to be left alone (20.0%), and the belief that no one can care better (8.6%).

In the course of taking care of their patients, majority (89.2%) spent ≤ 1000 naira per day on expenses related to themselves (Table 3). The mean and median amount caregivers spent on themselves was found to be 803.94 naira and 600 naira respectively. Among those employed, about two thirds (63.2%) reported loss of income while caregiving. When asked to quantify the daily income lost, about four fifths (81.7%) reportedly lost an average of ≤ 1,000 naira daily. The mean and median average daily loss was 1115.7 naira and the median amount was 3000.0 naira.

**Table 3:** Caregivers’ Finances and Sources of Support

| **Variables** | **Frequency (%)** |
| --- | --- |
| **Estimated amount spent daily on self** |  |
| ≤ 1000 | 357 (89.2) |
| > 1000 | 43 (10.8) |
| Mean (Median) | 803.94 (600.0) |
| **Loss of income while caregiving (n = 277)** |  |
| Yes | 175 (63.2%) |
| No | 102 (36.8%) |
| **Estimated amount loss daily (n=175)** |  |
| ≤ 1000 | 143 (81.7) |
| > 1000 | 32 (18.3) |
| Mean (Median) | 1115.7 (3000.0) |
| **Received financial support for self** |  |
| Yes | 44 (11.0) |
| No | 356 (89.0) |
| **Sources of financial support for self(n=44)** |  |
| Children | 5 (11.4) |
| Other relatives | 23 (52.3) |
| Friends | 10 (22.7) |
| Parents | 6 (13.6) |
| **Estimated amount gotten for self (n=44)** |  |
| ≤ 5000 | 21 (47.7) |
| > 5000 | 23 (52.3) |
| Mean (Median) | 21931.8 (6500.0) |
| **Estimated amount spent daily on patient** |  |
| < 5000 | 183 (45.7) |
| ≥ 5000 | 217 (54.3) |
| Mean (Median) | 8835.14 (5000.0) |
| **Most frequent needs of patient** |  |
| Food | 57 (14.3) |
| Drugs | 337 (84.2) |
| Investigations | 6 (1.5) |
| **Provide financial support to patient** |  |
| Yes | 203 (50.8) |
| No | 197 (49.2) |
| **Sources of financial support for patient** | n=203 |
| Children | 7 (3.4) |
| Other relatives | 125 (61.6) |
| Friends | 35 (17.2) |
| Parents | 11 (5.4) |
| Association/Office | 8 (3.9) |
| Church members | 12 (5.9) |
| Others | 5 (2.5) |
| **Estimated amount gotten for patient** |  |
| ≤ 5000 | 17 (8.4) |
| > 5000 | 186 (91.6) |
| Mean (Median) | 73200 (40000.0) |

Only one tenth (11.0%) received financial support for themselves while caregiving. The commonest sources of support were other relatives (52.3%), friends (22.7%), parents (13.6%) and children 5 (11.4%). Slightly more than half (52.3%) had gotten >5000 naira daily from these sources and the mean and median amount was estimated to be 21931.8 naira and 6500.0 naira respectively. More than half (54.3%) of the respondents spent < 5000 naira daily on their patients. The mean and median amount spent daily was estimated to be 8835.14 naira and 5000 naira correspondingly. Drugs, food and investigations accounted for 84.2%, 14.3% and 1.5% of the patient’s needs respectively.

Financial support for the patient was also gotten from other relatives (61.6%), friends (17.2%), church members (5.9%), parents (5.4%) and children (3.4%). Almost all (91.6%) of the respondents had gotten <5000 naira for the care of their patient; the mean and median amount gotten were 73,200 and 40,000 naira respectively.

## Discussion

Caregiving cuts across most sociodemographic characteristics though some sects may be involved than others. In this study, the mean age of caregivers was about 38 years; this is similar to findings from South West Nigeria[13] and South South Nigeria.[11] This suggests that majority of caregivers in Nigeria are within the age group of the active labour force. The study also found a preponderance of the female gender among caregivers. This corroborates with the known social construct in which women are central to caregiving within the family setting and also in health care settings.[1,14] Most of the respondents were Yoruba and also Muslims. This may be related to the fact that majority of the inhabitants in the study area are Yoruba and practice Islam.[15]

This study found that almost all the caregivers supported their patients physically (running errands and assisting with activities of daily living) and financially while on hospital admission in a tertiary health institution in North Central Nigeria. This is similar to findings from other parts of Nigeria[11,13,16] and other developing countries like India[1] and Iran.[8] Common to these countries is the shortage of healthcare workers[17,18] and the existing social cohesive system[19,20] where informal caregivers play a vital supportive role to their patients during hospital admission.

Almost all the caregivers were relatives of the patients, with most of them being first degree relatives. This is similar to findings from other studies in Nigeria[11,13,21]and in India.[1]A probable reason for this is the cultural values and norms of caring within the family system in these settings. More than two thirds of the caregivers were employed and out of these, most of them were self-employed. This is in congruence with findings from South West Nigeria[13] but incongruent with findings from South Nigeria where majority of caregivers were unemployed or retired.[11] Being self-employed may infer that they are more in control and be more flexible of their time and can afford to stay with their patients in the hospital during work hours compared to those who are employees and are accountable to their employers. Findings from the study show an array of support types rendered by caregivers to their patients while on admission. Running of errands, feeding patients, serving medications, helping with defecation and bathing patients were the commonest forms of care reported. These errands were in form of buying medications, supplying non-medical needs, paying for services and running errands to and from the laboratory. This finding is not unique to the study area, as studies conducted in Nigeria and other developing countries have found that informal caregivers perform nursing duties and also run other errands in the hospital.[1,8,11] This may be due to the fact that healthcare workers may be overwhelmed. Thus, caregivers are needed to support with “non-technical” duties.

More than four fifths of the caregivers perceived staying around their patients in the hospital to be stressful. This is similar to findings to a previous study conducted in Nigeria where caregivers continued to care for their patients in spite of the perception that caregiving is a source of burden.[11]Closely linked to this is the fact that less than one fifths of them felt their patients would be well cared for in their absence. In addition, there was also a common perception that their patients will not be well cared for if they were not around due to inadequate staffing, the patient being too young to be left alone, the perception that no one can care better than them and the non-challant attitude of healthcare workers in the hospital. Olowookere et al found that about two fifths of caregivers were not satisfied with the hospital care received by their patient.[13]

Another important finding was this study was the gap in communication between caregivers and health workers. About one tenth of the caregivers did not know the type of ailment their patient was being managed for. Similar studies have also identified communication gap between healthcare professionals and caregivers in developed and developing countries.[1,22] Caregivers appreciably contribute a supportive role in patient management, and more so during transition of care following hospital discharge.[23] Good communication with healthcare workers is associated with a reduced risk of readmission to the hospital.[24] It is therefore imperative that healthcare workers involve carers, and communicate the nature of illness, management process and follow up care to achieve optimal patient-related outcomes.

The study also elucidates the financial burden experienced by caregivers. Implications from this study suggest that caregivers lose daily income while they are away from their workplace, giving care to their patients in the hospital. In addition, some spend their money on patients and also have to spend money on themselves while in the hospital. Majority of them do not receive financial support from other sources during this period while they experience increased financial expenses and also a decrease in their income. Similarly, other studies conducted in Nigeria among informal caregivers have revealed that financial burden as a major challenge when caring for their loves ones.[11,16] In a study conducted in Nigeria among caregivers, all of the respondents claimed that the medical bills of their patient was paid out of pocket.[13]The financial burden faced by these caregivers further highlights the bane of the limited financial risk protection mechanisms for healthcare services in the country. Nigeria is a country where less than 5% and only 3% of people in the formal and informal sectors respectively have health insurance.[25] Health insurance can go a long way in reducing the financial burden faced by patients and their caregivers.

## Limitations

This was a cross sectional study which was conducted in one health facility. The results from this study might not be generalizable to the entire country. Factors associated with the support types and the extent of financial burden experienced by caregivers were not identified. Further research will be needed to identify these factors.

## Conclusion

This study provides valuable information on the extent of physical and financial support, and other challenges experienced by caregivers while staying with their patients on hospital admission. Findings from this study underpin the need for policy makers, hospital administrators and other stakeholders to consider the role caregivers play in patient management and also institute processes to ease the burden that they face. The challenges caregivers experience can be eased off by the simplification of payment processes, and assigning a dedicated staff for the submission of laboratory samples, retrieval of results, and the purchase of medical supplies. These will bring about changes that will results in a more caregiver friendly environment. Outcomes from this study also highlights the need for effective communication between healthcare professionals and caregivers in the management process of their patients. Furthermore, the financial burden experienced by caregivers reinforces the need to encourage more Nigerians to enrol in a health insurance scheme.

### What is known on this topic?

- Informal caregivers play a role in patient management, particularly in middle- and low-income countries
- Caregiver burden has been linked to a negative impact on their physical, psychological and emotional health

### What this study adds

- Informal caregivers experience significant physical and financial burden
- There is communication gap between healthcare workers and the informal caregivers

## Data Availability

All data produced in the present study are available upon reasonable request to the authors

## Author’s Contribution

All authors made significant contribution to the conception and design, acquisition of data or analysis and interpretation of data. OWA wrote the first draft of the manuscript. All authors made substantial intellectual contribution to all drafts of the manuscript, and approved the final draft.

